# Impact of weather indicators on the COVID-19 outbreak: A multi-state study in India

**DOI:** 10.1101/2020.06.14.20130666

**Authors:** Kuldeep Singh, Aryan Agarwal

**Affiliations:** Department of Electronics and Communication Engineering, Malaviya National Institute of Technology, Jaipur, India

**Keywords:** COVID-19, Correlation, Temperature, Humidity, India

## Abstract

The present study examines the impact of weather indicators on the COVID-19 outbreak in the majorly affected states of India. In this study, we hypothesize that the weather indicators could significantly influence the impact of the corona virus. The Kendall and Spearman rank correlation tests were chosen to conduct the statistical analysis. In this regard, we compiled a daily dataset including confirmed case counts, Recovered case counts, Deceased cases, Average Temperature, Maximum Relative Humidity, Maximum Wind Speed for six most affected states of India during the period of March 25, 2020 to April 24, 2020. We investigated that the average Humidity and Average Temperature seven days ago play a significant role in the recovery of coronavirus cases. The rise in average temperature will improve the recovery rate in the days to come. The cities with very high humidity levels or dry weather conditions have high probabilities of recovery from COVID-19. The findings of this research will help the policymakers to identify risky geographic areas and enforce timely preventive measures.

## 1. Introduction

Coronavirus disease 2019 (COVID-19) is an infectious disease which initially detected in Wuhan, China, has now spread all over the world, and if not well dealt, it could even lead to the worldwide economic crisis. This virus exhibits high human-to-human transmissibility, that is the reason it has spread all across the world in a very short span of time. The World Health Organization (WHO) reported 2,626,321 COVID-19 confirmed cases and 181,938 deaths worldwide until April 24, 2020 [1]. The deadly virus has affected more than 210 countries had been affected where United States (US) alone contributed approximately one-fourth of the total cases. Other major countries having a significant impact are Spain, Italy, Iran, France, Germany, UK, Turkey, China, and many more. The situation in India has started worsening day by day. The first COVID-19 case in India was reported on January 30, 2020 and as on April 24, 2020 India has 23077 COVID-19 confirmed cases, including 718 deaths [1]. As per WHO, India has not entered into the state of the community transmission, still it has clusters of cases. India had enforced nationwide lockdown from March 25, 2020 to April 14, 2020 and further extended it till May 3, 2020. The lockdown might have lowered the transmission rate of the COVID-19 pandemic. Figure 1 shows the overall picture of confirmed cases of COVID-19 across the states of India. Many geographical, social, political, and environmental factors might influence the impact of this deadly virus.

**Fig 1.**
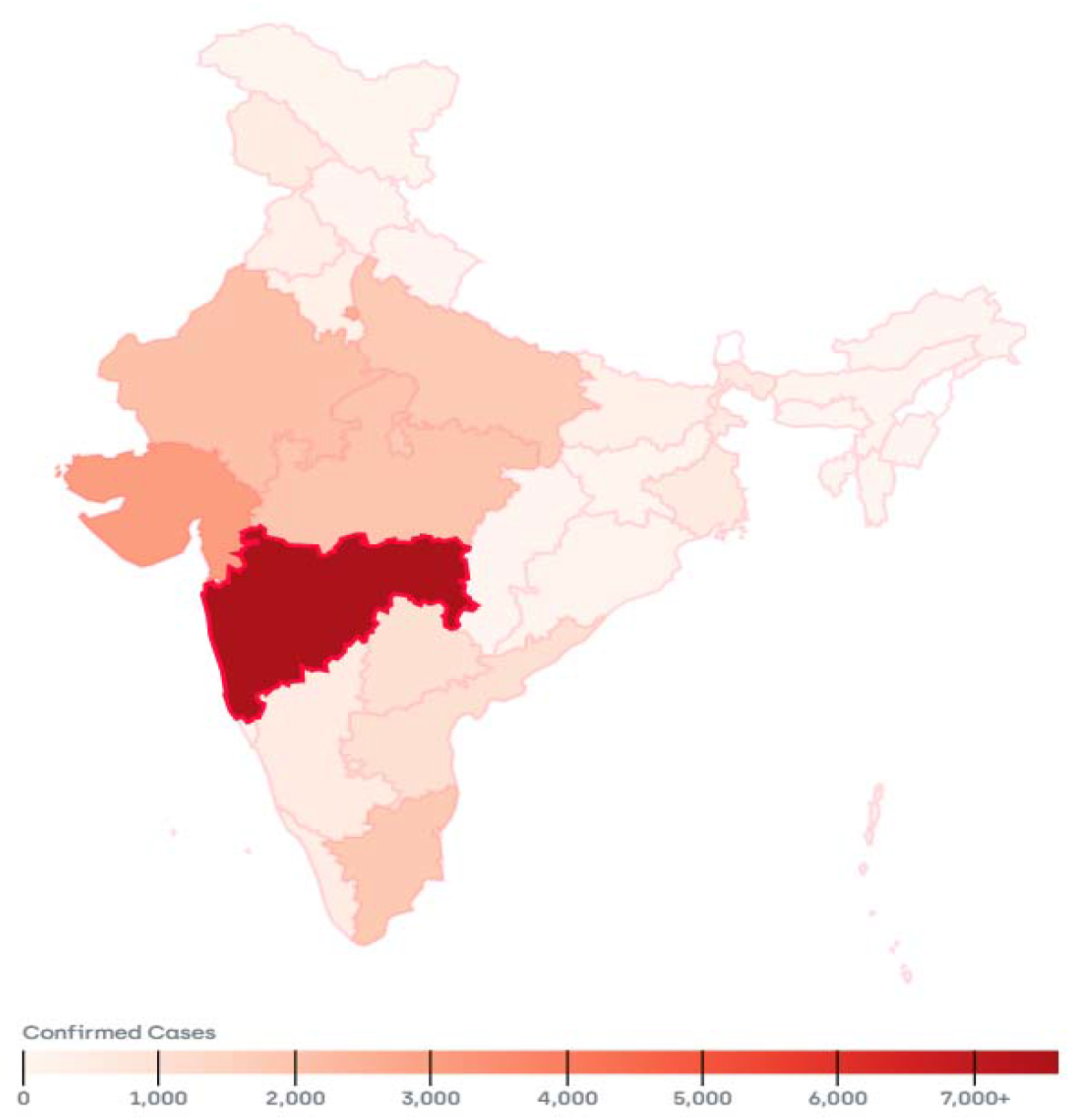
Outbreak of COVID-19 in India (Source: https://www.covid19india.org/)

Earlier studies have suggested that the meteorological parameters might be active factors in the transmission of viruses and disease emergence. Yuan et al. investigated that the peak spread of SARS occurred in a particular range of Temperature, Relative Humidity, and Wind Velocity [2]. Variations of absolute Humidity correlate with the onset and seasonal cycle of influenza viral in the US. [3]. Recently, various studies have been conducted to analyze the impact of weather conditions on the spread and effect of COVID-19. We have summarized the outcome of these studies conducted worldwide in the Table1.

**Table 1:**
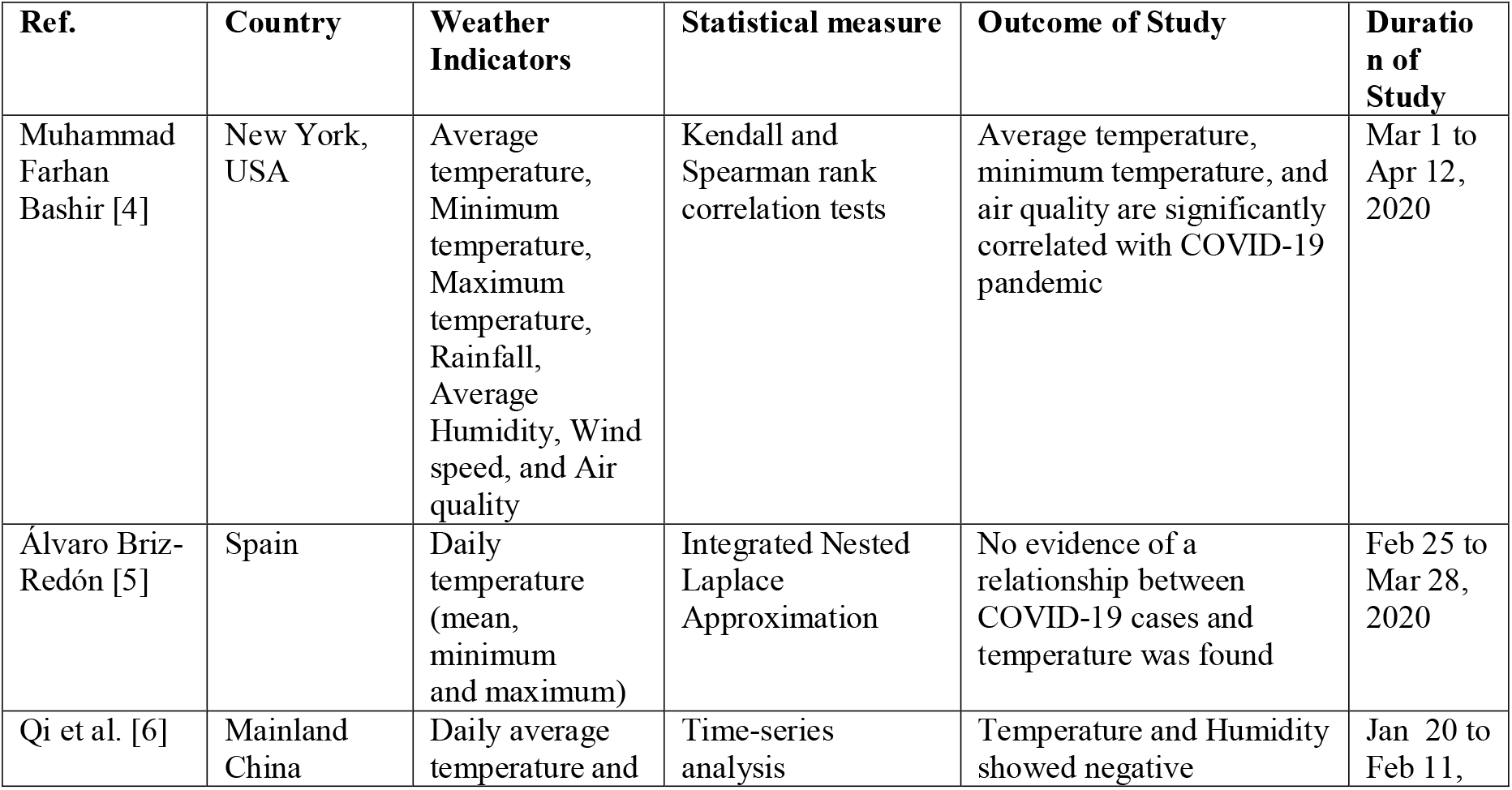
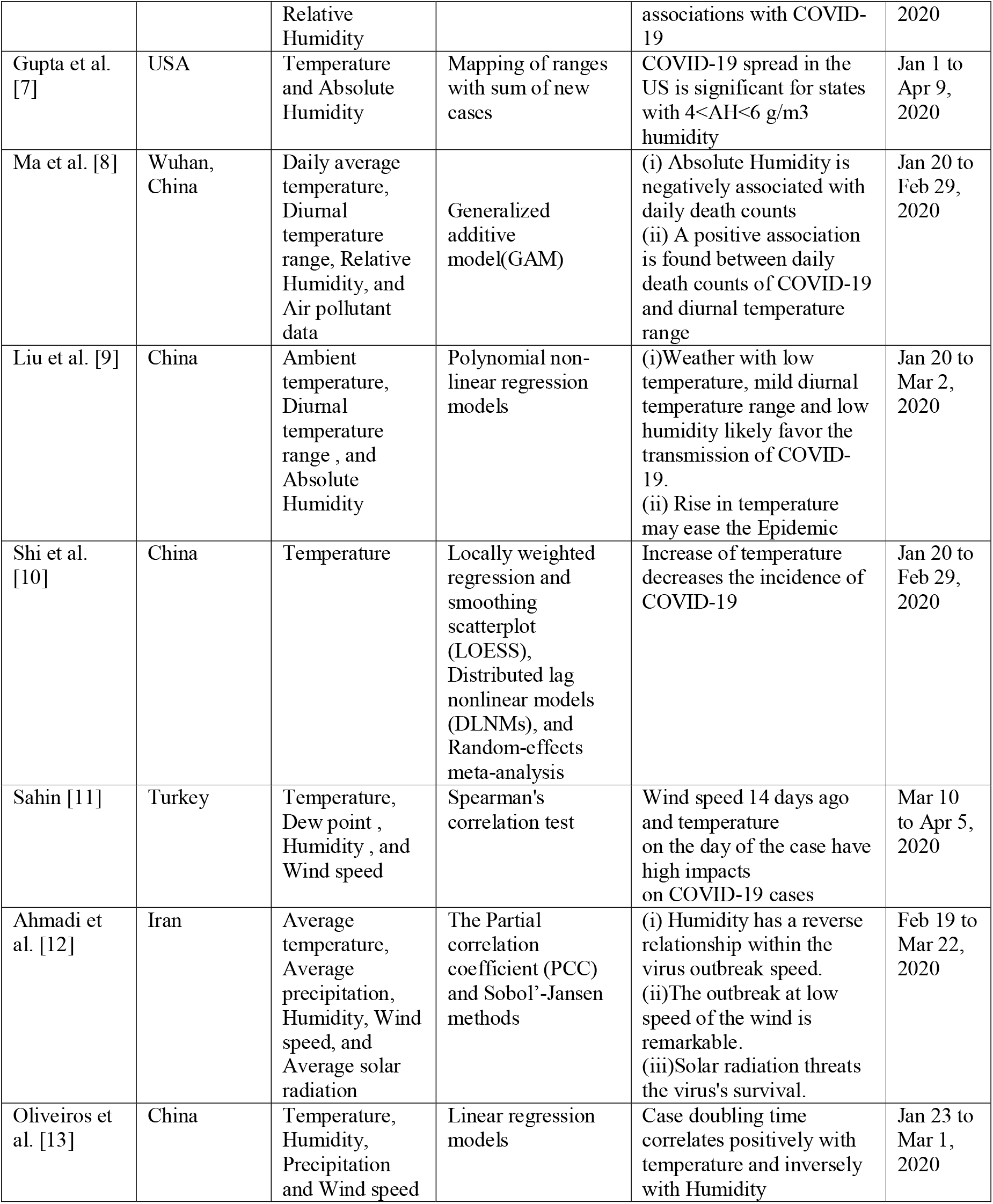
Recent studies on the impact of weather condition on COVID-19 Spread

The independent effect of weather indicators on the transmission of COVID-19 has not been studied systemically in the Indian context. The main objective of this study is to investigate the association between weather indicators and the COVID-19 outbreak in India. To the best of our knowledge, this is the first study to explore the effects of weather indicators on COVID-19 outbreak for India. Since every state has a different geographical and political environment, we have considered different states of India rather than on state or city. The primary weather indicators i.e. temperature (°C), wind speed (mph), and Humidity (%) are considered as independent variables for finding the correlation with affected cases of COVID-19. The majority of the related recent studies have considered the metrological indicators of the same day, which could present a false picture in terms of the correlation with COVID-19 cases. Since the incubation period of the COVID-19 virus varies from 1 day to 14 days, we have evaluated one-week old weather indicators.

## 2. Materials and Method

### 2.1 COVID-19 Data

The data of COVID-19 cases of India corresponding to the six most impacted states Maharashtra, Delhi, Rajasthan, Gujarat, Tamilnadu, and Madhya Pradesh was retrieved from a publicly available repository and accessible through this link: https://www.covid19india.org/. This is a volunteer-driven, crowd-sourced database being collected and homogenized from multiple official and private web sources.

### 2.2 Weather Data

The weather data, including Temperature (Max, Min, Average), Humidity (Max, Min, Average) and Wind Speed (Max, Min, Average) of cities contributing the highest number of cases in the six most impacted states i.e. Maharashtra(Mumbai), Delhi, Rajasthan(Jaipur), Gujarat (Ahmedabad), Madhya Pradesh (Indore), and Tamilnadu (Chennai) were retrieved from Weather Underground (https://www.wunderground.com/). In this study, the correlation of all these weather indicators was computed; however, for the sake of better clarity, only the weather parameters showing significant results were shown in this paper.

Figure 2 shows a comprehensive combo chart showing the weather indicators (Maximum relative Humidity, Average Wind speed, and Average Temperature) and the COVID-19 statistical indicators of the chosen six states in one glance.

**Fig 2.**
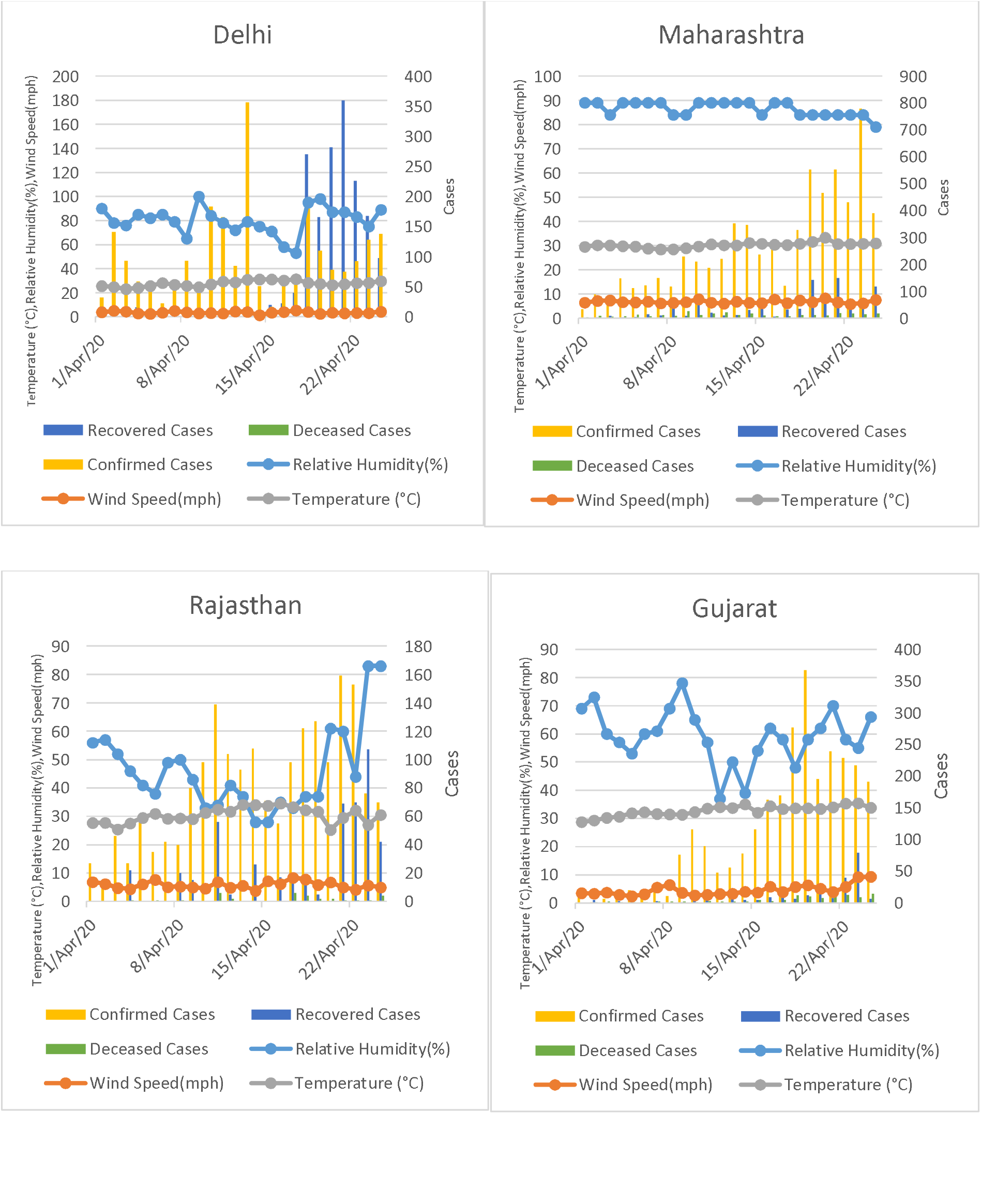
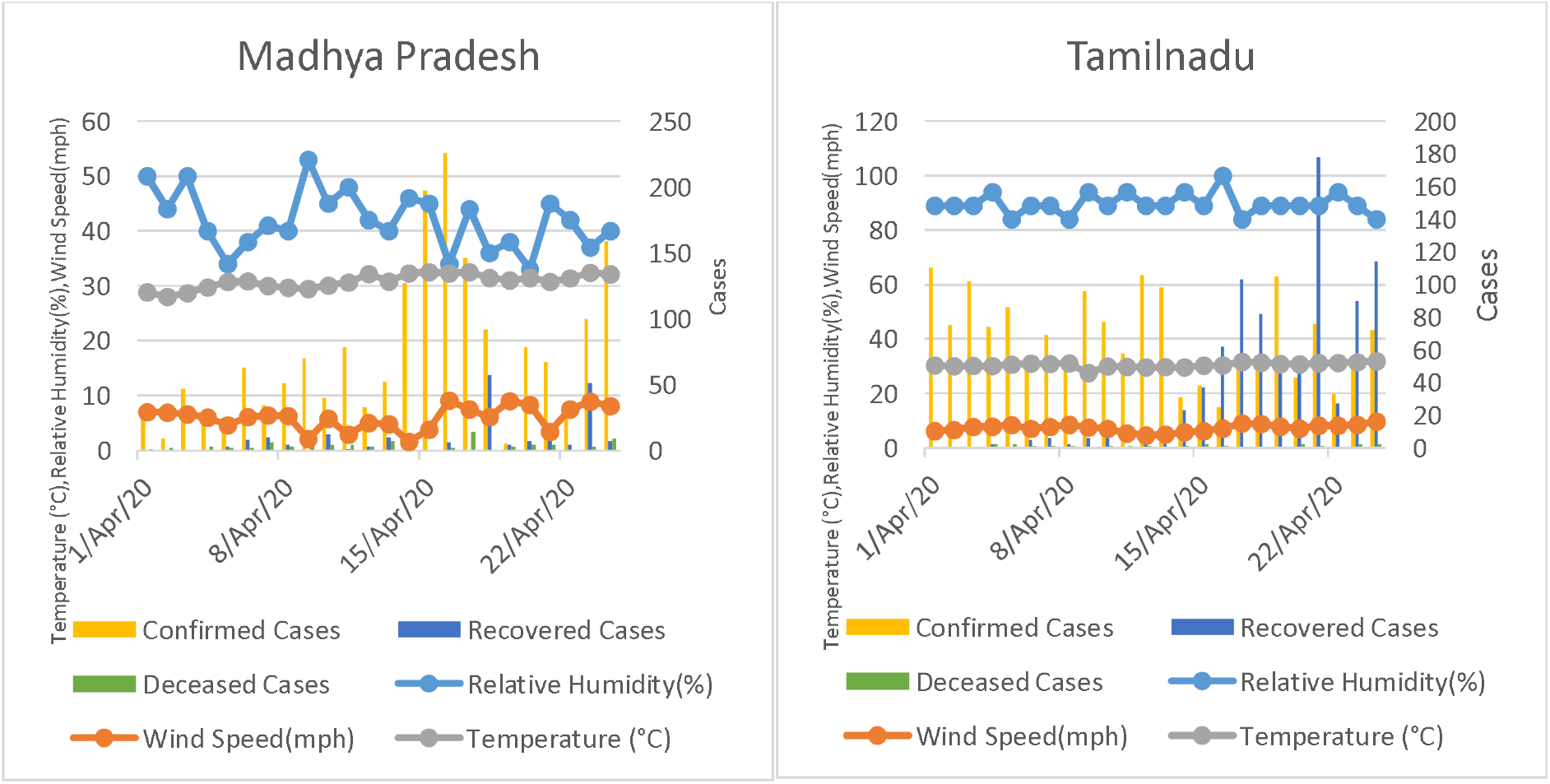
COVID-19 case statistics and weather indicators of six states during the period of April 1, 2020 to April 24, 2020.

### 2.3 Statistical analysis

The weather and COVID-19 data collected are not normally distributed; therefore Kendall [14] and Spearman rank correlation [15] tests are considered in this study to investigate the correlation between weather indicators and COVID-19 cases. Both of these methods are accepted measures of non-parametric rank correlations. A null hypothesis corresponding to each weather indicator is formulated that there is no association between individual indicators and the three case statistics considered in this study. For all the hypothesis testing, statistical significance was set at p-value < 0.05.

## 3. Results and Discussion

Table 2 highlights the Kendall and Spearman’s correlation coefficients corresponding to six weather indicators among the confirmed, recovered, and deceased cases for the six states. The bold values show moderate or strong correlations among the variables with 5% significance level.

**Table 2:** Correlation coefficients between weather indicators and COVID-19 cases

| State | Weather Indicators | Kendall Correlation Coefficient |  |  | Spearman Correlation Coefficient |  |  |
| --- | --- | --- | --- | --- | --- | --- | --- |
|  |  | Confirmed Cases | Recovered Cases | Deceased | Confirmed Cases | Recovered Cases | Deceased |
| Maharashtra | Average Temperature (Same Day) | **0.525** | **0.419** | 0.168 | **0.727** | **0.593** | 0.277 |
|  | Max. Relative Humidity (%) | **-0.405** | **-0.532** | -0.214 | **-0.508** | **-0.639** | -0.258 |
|  | Max. Wind Speed (mph) | 0.256 | 0.184 | 0.253 | 0.327 | 0.262 | 0.333 |
|  | Average Temperature (7 Days ago) | 0.235 | 0.239 | 0.292 | 0.341 | 0.347 | **0.410** |
|  | Average Relative Humidity (7 Days ago) | **0.458** | **0.317** | 0.133 | **0.628** | **0.437** | 0.228 |
| Delhi | Average Temperature (Same Day) | 0.059 | 0.087 | **0.359** | 0.077 | 0.185 | **0.508** |
|  | Max. Relative Humidity (%) | 0.063 | 0.155 | -0.280 | 0.050 | 0.253 | -0.363 |
|  | Max. Wind Speed (mph) | 0.064 | 0.041 | 0.009 | 0.083 | 0.102 | 0.027 |
|  | Average Temperature (7 Days ago) | 0.143 | **0.558** | 0.020 | 0.233 | **0.728** | 0.053 |
|  | Average Relative Humidity (7 Days ago) | -0.201 | **-0.402** | -0.113 | -0.299 | **-0.591** | -0.192 |
| Rajasthan | Average Temperature (Same Day) | 0.289 | 0.124 | 0.106 | **0.466** | 0.163 | 0.091 |
|  | Max. Relative Humidity (%) | -0.229 | 0.056 | 0.047 | -0.321 | 0.113 | 0.075 |
|  | Max. Wind Speed (mph) | -0.082 | -0.055 | 0.323 | -0.101 | -0.118 | 0.381 |
|  | Average Temperature (7 Days ago) | **0.482** | **0.394** | **0.361** | **0.635** | **0.501** | **0.517** |
|  | Average Relative Humidity (7 Days ago) | **-0.547** | **-0.354** | **-0.318** | **-0.721** | **-0.459** | **-0.441** |
| Gujarat | Average Temperature (Same Day) | **0.536** | **0.448** | **0.511** | **0.724** | **0.611** | **0.661** |
|  | Max. Relative Humidity (%) | -0.099 | -0.132 | -0.047 | -0.159 | -0.202 | -0.084 |
|  | Max. Wind Speed (mph) | 0.121 | 0.124 | **0.324** | 0.186 | 0.152 | 0.397 |
|  | Average Temperature (7 Days ago) | **0.696** | **0.455** | **0.623** | **0.868** | **0.637** | **0.806** |
|  | Average Relative Humidity (7 Days ago) | **-0.489** | **-0.303** | **-0.345** | **-0.635** | **-0.426** | **-0.454** |
| Madhya Pradesh | Average Temperature (Same Day) | **0.486** | 0.255 | 0.023 | **0.657** | 0.341 | 0.042 |
|  | Max. Relative Humidity (%) | -0.026 | **-0.318** | 0.068 | -0.036 | **-0.424** | 0.080 |
|  | Max. Wind Speed (mph) | -0.049 | 0.047 | -0.138 | -0.034 | 0.050 | -0.154 |
|  | Average Temperature (7 Days ago) | 0.212 | **0.406** | 0.246 | 0.320 | **0.555** | 0.314 |
|  | Average Relative Humidity (7 Days ago) | -0.222 | **-0.319** | -0.233 | -0.299 | **-0.464** | -0.350 |
| Tamilnadu | Average Temperature (Same Day) | -0.193 | **0.405** | 0.084 | -0.324 | **0.554** | 0.115 |
|  | Max. Relative Humidity (%) | -0.199 | -0.071 | -0.106 | -0.240 | -0.091 | -0.123 |
|  | Max. Wind Speed (mph) | -0.121 | 0.170 | -0.063 | -0.157 | 0.209 | -0.086 |
|  | Average Temperature (7 Days ago) | -0.091 | 0.217 | 0.128 | -0.144 | 0.308 | 0.137 |
|  | Average Relative Humidity (7 Days ago) | -0.127 | **0.463** | 0.044 | -0.186 | **0.656** | 0.056 |

The significant findings of this study are the strong correlation between average temperature 7 days ago and average relative Humidity 7 days ago with the recovered cases. The average temperature 7 days ago has shown moderate correlation (0.394 < r < 0.728) with recovered cases in four states i.e. Delhi, Rajasthan, Gujarat, and Madhya Pradesh for both Kendall and Spearman correlation test. The rise in average temperature might improve the recovery rate in the days to come. In Rajasthan and Gujarat, average temperature 7 days ago parameter is also associated with new confirmed cases and deceased cases. Similarly, the average temperature same day has shown some associations with confirmed and recovered cases as posited by [16]. These outcomes might be due to a certain profile of the temperature in these states. [17] also reported similar findings that warm weather would play an important role in suppressing the virus.

In this study, it has been analyzed that the states having different humidity profiles exhibit different recovery response from COVID-19. The results show that average relative Humidity 7 days ago has a conclusive association with recovered cases in all the states. The two states Tamilnadu and Maharashtra, where the average relative Humidity is in range ∼ 65-85 % are positively correlated with recovered cases, which implies that recovery chances are higher in the humid environment. However, the other states where humidity levels are comparatively low and range approximately between 20% to 60% has a negative correlation between relative Humidity 7 days ago with the recovered cases. It can be concluded that the cities with very high humidity levels or very low humidity levels have high probabilities of recovery from COVID-19. By and large, the maximum relative Humidity on the same day doesn’t exhibit a significant association with the COVID-19 cases in the duration of this study, which contradicts the study in Iran [12]. In the time interval of this study, we observed that the wind speed does not affect the viral spread or recovery in all the states considered.

This study has shown evidence of weather indicators correlation with COVID-19 cases; however, there are various limitations under which this study has been conducted. The variables such as lockdown measures, people’s individual immunity, migration index, and other climate indicators can impact the results presented in this study.

## 4. Conclusions

The weather indicators can play a crucial role in the fight against coronavirus. To the best of our knowledge, this is the first study to investigate the impact of weather indicators on COVID-19 incidences. This study investigates that average temperature and average relative Humidity seven days ago are significantly correlated with the COVID-19 outbreak and will be useful in suppressing COVID-19. In conclusion, weather indicators influence the COVID-19 pandemic, potentially hot, and humid environment that can help in the recovery of the infected patients. The present study can be further enhanced by including other parameters such as demographic variations, healthcare infrastructure, and social policies like lockdowns to provide better insight into the fight against COVID-19.

## Data Availability

The data that support the findings of this study are available from OpenAQ platform and Weather Underground. These data were derived from the following resources available in the public domain: https://www.covid19india.org/, https://openaq.org and https://www.wunderground.com/

https://www.wunderground.com/

## Acknowledgments

This research was supported partially by the research grant scheme of TEQIP-III, MNIT Jaipur.

## Notes

### Competing Interest Statement

The authors have declared no competing interest.

### Author Declarations

Ethics committee approvals not required for this study

## Reference

[1] WHO, “Coronavirus disease 2019 (COVID-19) Situation Report – 95,” World Health Organization, 2020.

[2] J. Yuan, H. Yun, W. Lan, W. Wang, S. G. Sullivan, S. Jia and A. H. Bittles, “A climatologic investigation of the SARS-CoV outbreak in Beijing, China,” American Journal of Infection Control, vol. 34, no. 4, pp. 234–236, 2006.

[3] J. Shaman, E. Goldstein and M. Lipsitch, “Absolute Humidity and Pandemic Versus Epidemic Influenza,” American Journal of Epidemiology, vol. 173, no. 2, p. 127–135, 2011.

[4] M. F. Bashir, B. Ma, B. B. Komal, M. A. Bashir, D. Tan and M. Bashir, “Correlation between climate indicators and COVID-19 pandemic in New York, USA,” Science of The Total Environment, vol. 728, 2020.

[5] Á. Briz-Redón and Á. Serrano-Aroca, “A spatio-temporal analysis for exploring the effect of temperature on COVID-19 early evolution in Spain,” Science of The Total Environment, 2020.

[6] H. Qi, S. Xiao, R. Shi, M. P. Ward, Y. Chen, W. Tu, Q. Su, W. Wang, X. Wang and Z. Zhang, “COVID-19 transmission in Mainland China is associated with temperature and humidity: A time-series analysis,” Science of The Total Environment, 2020.

[7] S. Gupta, G. S. Raghuwanshi and A. Chand, “Effect of weather on COVID-19 spread in the US: A prediction model for India in 2020,” Science of The Total Environment, vol. 728, 2020.

[8] Y. Ma, Y. Zhao, J. Liu, X. He, B. Wang, S. Fu, J. Yan, J. Niu, J. Zhou and B. Luo, “Effects of temperature variation and humidity on the death of COVID-19 in Wuhan, China,” Science of The Total Environment, vol. 724, 2020.

[9] J. Liu, J. Zhou, J. Yao, X. Zhang, L. Li, X. Xu, X. He, B. Wang, S. Fu, T. Niu, J. Yan, Y. Shi, X. Ren, J. Niu, W. Zhu, S. Li, B. Luo and K. Zhang, “Impact of meteorological factors on the COVID-19 transmission: A multi-city study in China,” Science of The Total Environment, vol. 726, 2020.

[10] P. Shi, Y. Dong, H. Yan, C. Zhao, X. Li, W. Liu, M. He, S. Tang and S. Xi, “Impact of temperature on the dynamics of the COVID-19 outbreak in China,” Science of The Total Environment, 2020.

[11] M. Şahin, “Impact of weather on COVID-19 pandemic in Turkey,” Science of the Total Environment, vol. 728, 2020.

[12] M. Ahmadi, A. Sharifi, S. Dorosti, S. J. Ghoushchi and N. Ghanbari, “Investigation of effective climatology parameters on COVID-19 outbreak in Iran,” Science of The Total Environment, 2020.

[13] B. Oliveiros, L. Caramelo, N. C. Ferreira and F. Caramelo, “Role of temperature and humidity in the modulation of the doubling time of COVID-19 cases,” medRxiv, 2020.

[14] M. Kendall, “A New Measure of Rank Correlation,” Biometrika, vol. 30, pp. 81-89, 1938.

[15] C. Spearman, “The proof and measurement of association between two things,” The American Journal of Psychology, vol. 15, no. 1, pp. 72–101, 1904.

[16] R. Tosepu, J. Gunawan, D. S. Effendy, L. O. A. I. Ahmad, H. Lestari, H. Bahar and P. Asfian, “Correlation between weather and Covid-19 pandemic in Jakarta, Indonesia,” Science of the Total Environment, vol. 725, 2020.

[17] Y. Wang, Y. Wang, Y. Chen and Q. Qin, “Unique epidemiological and clinical features of the emerging 2019 novel coronavirus pneumonia (COVID-19) implicate special control measures,” Journal of medical virology, 2020.

